# ACCURACY OF COMPUTER-ASSISTED DETECTION IN SCREENING PEOPLE WITH DIABETES MELLITUS FOR ACTIVE TUBERCULOSIS: A SYSTEMATIC REVIEW

**DOI:** 10.1101/2024.12.10.24318764

**Authors:** Emoru Daniel Reagan, Lucy Elauteri Mrema, Nyanda Elias Ntinginya, Irene Andia Biraro, Reinout van Crevel, Julia A Critchley

**Affiliations:** Makerere University, College of Health Sciences, Department of Internal Medicine, Uganda; City St George’s, University of London, School of Health & Medical Sciences, Population Health Research Institute, London SW17 0RE, United Kingdom; Radbound University Medical Centers, Nijmegen, Netherlands; National Institute for Medical Research, Mbeya Tanzania

**Keywords:** diabetes mellitus, tuberculosis, CAD, systematic review

## Abstract

**Objectives:** Diabetes mellitus (DM) significantly increases the risk of tuberculosis (TB), and active TB screening of people with DM has been advocated. This systematic review aimed to evaluate the accuracy of computer-assisted detection (CAD) for identifying pulmonary TB among people living with DM.

**Methods:** Medline, Embase, Scopus, Global Health and Web of science were searched from January 2010 to May 2024 supplemented with grey literature (Conference abstracts, Trial registries, MedRxiv.org). Studies evaluating CAD accuracy for identifying TB in populations living with diabetes were included. Two researchers independently assessed titles, abstracts, full texts, extracted data and assessed the risk of bias. Due to heterogeneity and a limited number of studies, a descriptive analysis was performed instead of statistical pooling. Forest plot and Summary Receiver Operating Curves (SROC) were generated using RevMan 5.4.

**Results:** Five eligible studies, all conducted in Asia between 2013 and 2023 were identified, including a total of 1879 individuals of whom 391 were diagnosed with TB. Four different Computer Assisted Detection (CAD) software algorithms were used. Sensitivities ranged from 0.73 (95%CI: 0.61-0.83) to 1.00 (95%CI:0.59-1.00), while specificities ranged from 0.60 (95%CI:0.53-0.67) to 0.88 (95%CI: 0.84-0.91). Area Under the receiver Operating Curve (AUC) values varied from 0.7 (95%CI: 0.68-0.75) to 0.9(95%CI: 0.91-0.96). False positive rates ranged from 0.24% to 30.5%, while false negative rates were 0-3.2%. The risk of bias assessment of the five studies was generally good to excellent.

**Conclusions:** CAD tools show promise in screening people living with diabetes for active TB, but data are scarce, and performance varies across settings.

## Introduction

Tuberculosis (TB) continues to pose a significant global health challenge, further exacerbated by the rising prevalence of diabetes mellitus (DM) which is a recognized risk factor for active TB(1). Individuals with DM are estimated to have a threefold increased risk of developing TB compared to those without DM(2,3). The International Diabetes Federation (IDF) estimated that in 2021, there were 537 million adults with DM, a number projected to rise to 783 million by 2045(4), rising the most in some regions where TB is still endemic such as sub-Saharan Africa.

WHO advocates TB screening for high-risk individuals in healthcare settings including people with DM, especially in countries where TB prevalence exceeds 100/100,000, where resources permit(5). International Union Against Tuberculosis and Lung disease/World Diabetes Forum guidelines (IUATLD/WDF) also recommend TB screening for newly diagnosed DM patients. Despite this guidance, practical constraints such as limited diagnostic resources, have hindered implementation(6).

However, advances in artificial intelligence (AI) have shown promise in enhancing the detection and management of health conditions including TB and DM complications(7).

In the field of diabetes management, computerised detection of complications is being increasingly adopted(8). A notable example is the implementation of AI-based retinopathy screening among people with diabetes in the UK(9). Computer assisted detection (CAD)tools, such as CAD4TB, qXR and LUNIT INSIGHT CXR utilise advanced machine-learning techniques to automatically analyse chest Xray (CXR) images and identify potential TB-associated abnormalities(10). All of these CAD tools operate on a similar principle: they analyse CXR images using machine learning algorithms and calculate an abnormality score, typically on a scale of 0-100, which represents the likelihood of TB-associated abnormalities being present in the image(11).

The potential of CAD in TB identification and screening has been recognized by global health authorities(10). Notably, both CAD4TB and qXR have been reviewed and approved by the World Health Organization (WHO) Guidelines Development Group for use in TB triage and screening in individuals aged 15 years and older, as of 2021(10)(12). This endorsement underscores the potential of these tools to significantly impact TB screening strategies globally.

However, despite these advancements, there is a notable gap in the literature regarding the application of CAD specifically for TB screening in people with DM. While systematic reviews have been conducted for other high-risk groups, such as those living with HIV in endemic African settings, the evidence might not necessarily apply to people with DM(12). The performance of CAD in this population might differ since DM is associated with obesity, heart failure, previous TB and pneumonia which can affect the appearance of chest radiographs. Also, TB patients with DM tend to be 10-20 years older than those without, and are therefore more likely to have other comorbidities which can affect the CXR interpretation e.g. lung cancer(13). Given the increased risk of TB in people with diabetes and the potential of CAD to enhance TB screening, there is a clear need to evaluate the accuracy and effectiveness of this technology specifically in people with DM(14). This review therefore aims to address this gap by systematically evaluating the available evidence on the accuracy of computer-assisted technology for active TB detection in populations living with DM.

## Methods

The protocol for this review was prespecified and published in PROSPERO (registration number (CRD42024523384) and we followed the PRISMA-DTA guidelines(15) for reporting the systematic review.

### Search Strategy

We searched Medline (via Ovid), Embase (via Ovid), Global Health (via Ovid), Web of Science and Scopus databases on 13/05/2024 for studies published between January 2010 and May 2024. Search strings included MESH, Keyword terms and synonyms for the words: ‘diabetes’, ‘tuberculosis’, and ‘Computer-aided detection’ (S1 Appendix). As this technology is relatively new, we supplemented with searches on 28/05/2024 of 1) International Union Against Tuberculosis and Lung Disease (IUATLD) conference abstracts (from 2018 to 2023), 2) the pre-print server medrxv; search was conducted from January 2010 to October 2021 because from November 2021 onwards, preprints were automatically captured by Embase databases. 3) Google Scholar (30/05/2024, first 20 pages).

### Eligibility Criteria

Studies were eligible for inclusion if they involved adult participants aged 15 years or older who had been diagnosed with DM. We included studies of general populations if they had been stratified by DM status. We also contacted authors to obtain data for people with DM if the study appeared to have stratified by DM status but had not reported results for this population group. The intervention needed to focus on computer-aided detection or screening methods, which could include artificial intelligence, machine learning, or deep learning approaches. Eligible studies (16-19) must have employed a reference standard involving microbiological confirmation of tuberculosis, such as sputum smear, culture, or Gene Xpert, which are recognised as reliable diagnostic methods by the WHO(20). Additionally, the studies had to report or have collected data to report diagnostic accuracy measures relevant to screening individuals with diabetes for active pulmonary tuberculosis in 2 by 2 table format. Case-control studies, reviews, editorials, commentaries, case reports or series, non-human studies and studies lacking full text availability were not considered for inclusion.

### Study selection and data extraction

Titles and abstracts of studies with the searches were screened independently by two researchers (EDR, JAC) with disagreement resolved by discussion. Of potentially relevant studies, full texts were obtained which were again screened by two researchers independently within Covidence (EDR, LEM). Any disagreements on eligibility were resolved through discussion or consultation with a third researcher (JAC).

From the studies included, the following data were extracted: study setting, period, inclusion and exclusion criteria, and target population. Available DM characteristics such as type of diabetes mellitus, diagnostic methods, duration, glycaemic control, and comorbidities were also documented. Details on TB screening modalities were noted, including the specific CAD technology used, symptom screening procedures, CXR reading methods, microbiological confirmation techniques, and the reference standard for active TB. Outcome measures were collected, including TB prevalence, sensitivity, specificity, predictive values, AUC, 95% confidence intervals, true positives (TP) and false negatives (FN).

Additional data on other screening tests, such as symptom screening and chest radiography, was also recorded if reported, including their sensitivity, specificity, predictive values and AUC. Details of study limitations, funding sources, conflicts of interest, and TB treatment details were noted. Diagnostic accuracy data for individuals without diabetes was also planned to be extracted for comparative purposes if reported.

### Risk of bias assessment

To assess the risk of bias of included studies, we applied QUADAS-2 tool; conducted by two researchers independently (EDR, ELM) with disagreements resolved by consultation with a third researcher (JAC) and results plotted using the Robvis tool(21).

### Data synthesis and statistical analysis

We constructed 2×2 contingency tables, using data on true positives (TP), false positives (FP), true negatives (TN), and false negatives (FN) for CAD software compared to the reference standard of microbiologically confirmed TB. We also estimated the TB prevalence and 95% CI in each included study, as this was rarely reported.

Coupled forest plots and SROC curves were generated in RevMan 5.4.1 to provide a visual representation of the studies’ sensitivities and specificities alongside their 95% CI, stratifying by software type. Meta-analysis using HSROC (Hierarchical summary receiver operating characteristic curve) model was not performed due to the small number of studies and heterogeneity between software types and results, suggesting that statistical pooling is not advisable(22).

## RESULTS

### Studies identified

A total 3097 citations were retrieved from the electronic database searches and grey literature, alongside 98 citations from reference list hand-searching. After removing 1461 duplicates (80 manually, 865 automatically by Covidence and 516 by other reasons), 1734 citations remained. Full texts for 7 studies were accessed, and 5 were included for final analysis (Figure 1).

**Figure 1:**
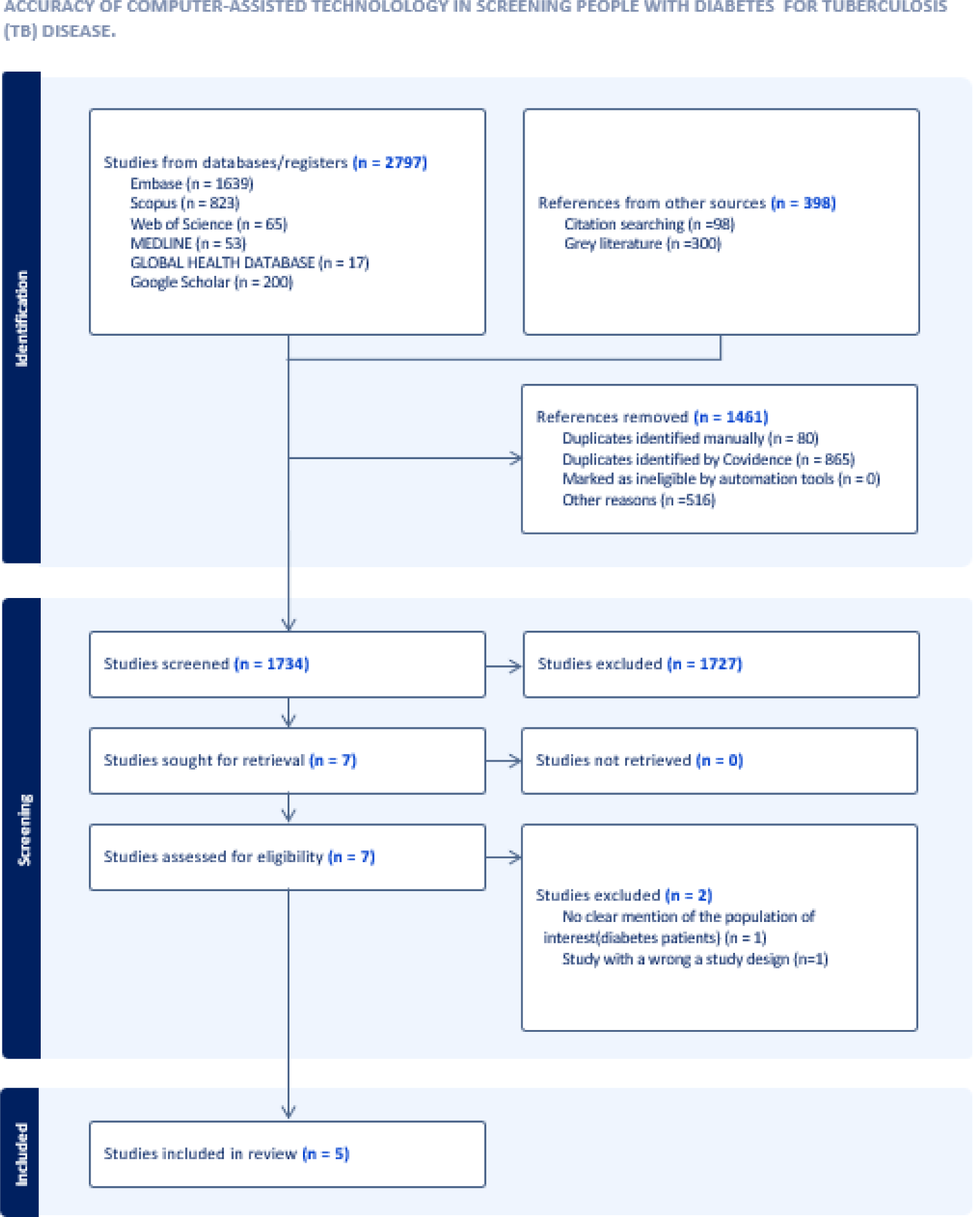
Prisma Flow Chart.

These five studies had included a total of 1879 individuals with DM, and newly diagnosed TB in 391 (19,17,18). All studies were cross sectional and conducted between 2013 and 2023 in Pakistan, India and Indonesia (24). Study participants had been enrolled in DM Clinics, TB treatment centres, diagnostic centres, outpatient clinics, community health centres, and mobile community screening units (Table 1).

**Table 1:** Descriptive Characteristics of the Included Studies.

| Author | Year of Study (Study Period) | Country | Age-group | Study design | *N | Index test | Index test threshold | Reference standard. |
| --- | --- | --- | --- | --- | --- | --- | --- | --- |
| Habib 2020 | July 2016 to April 2017 | Pakistan | ≥15 years | Cross-sectional | 694 | CAD4TB (version 3.07) | Between 50 and 90 | Xpert MTB/RIF |
| Khan 2020 | March 10, 2017, to July 13, 2018 | Pakistan | ≥15 years | Cross-sectional | 270 | 1.CAD4TB (version 6.0)<br>2. qXR (version 2.0) | 1. qXRv2, prespecified by manufacturer.<br>(not stated)<br>2. For CAD4TBv6, data-derived threshold of 57. | Culture-confirmed pulmonary tuberculosis. |
| Koesoemadinata 2018 | December 2013 to February 2015 | Indonesia | ≥ 18 years | Cross-sectional | 346 | CAD4TB (version 5.0) | Between 35 and 80, though a threshold score of 65 was chosen for the analysis. | Microbiological confirmation (Culture confirmed PTB or Xpert MTB/RIF |
| Nath 2024 | January 2018 to November 2023 | India | ≥ 15 years | Cross-sectional | 299 | DecXpert (Version 1.1) | It was not explicitly stated out in the article | Gene Xpert |
| Tavaziva 2022 | March 10, 2017, to July 13, 2018 | Pakistan | ≥15 years | Cross-sectional | 270 | LUNIT INSIGHT (version 3.1.0.0) | Manufacturer recommended thresholds of 15,30 and 45. Threshold of 30 was chosen. | Culture-confirmed Pulmonary Tuberculosis |

\*N-Represents the total number of diabetes patients who underwent automated chest Xray analysis.

Review of the included studies revealed limited information regarding specific DM characteristics. The Indonesian study(23)-Bandung Cohort did specify that most of the included participants had type 2 Diabetes whereas none of the other studies specified the type of diabetes as either type 1 or type 2(25). The Bandung cohort also showed varied diabetes duration, with the majority (38.4%) having diabetes for 1-5 years, followed by 35.4% for 6-15 years, while 17.8% had duration <1 year and 8.4% ≥15 years. Glycaemic control was distributed across categories, with 29.0% having HBA1c between 6.5-8.0%, 27.9%>10%, 25.3% between 8.1-10%, and 17.7%<6.5%(25). Finally, the Bandung cohort also did report data on smoking and alcohol use among its participants whereas the rest of the other studies did not report data on glycaemic control metrics, duration of diabetes, alcohol or smoking use given that these are important risk factors for TB.

Diabetes diagnosis varied between studies and was based on history of DM(18), single random blood glucose with history of levels >11.1mmol/l(16,18). Similarly, one study involved an extensive bidirectional screening program for TB and DM, including individuals with both known diabetes and those newly diagnosed with single random blood glucose levels >11.1mmol/l(16). One study enrolled adults ≥18yrs accessing diabetes care from the endocrine outpatient clinic, some were newly diagnosed(23). However, two of the studies did not specify how diabetes was diagnosed in their participants(17,19).

HIV status which increases TB risk and affects radiographic abnormalities in TB was reported in one study with 0.05% testing positive(18). One study reported comorbidities like Hypertension, Chronic lung and liver disease, Malignancy, Coronary artery disease and Chronic kidney disease(17). The same also study reported relevant use of co-medication like metformin, Insulin and other oral DM drugs(25) (which has been associated with less TB disease TB(26) or immunosuppressive drugs (which increase TB risk and may reduce extent of X-ray abnormalities)(27).

### Accuracy of CAD systems for TB diagnosis in people with DM

Five studies employed different CAD software with varying versions and thresholds [(16– 19,23)]. Three studies used CAD4TB software (two with different versions and one using both CAD4TB and qXR) (16,18,23), the fourth study used LUNIT INSIGHT CXR(19), and the fifth study used DecXpert(17). The CAD thresholds varied across studies; one study used CAD4TB thresholds between 50 and 90(16). Another study employed CAD4TB with a data derived threshold of 57 and qXR with a manufacturer specified threshold (not detailed in the article)(18). A third study utilized CAD4TB thresholds between 35 and 80, with a chosen threshold of 65 for analysis(23). The fourth study used manufacturer recommended LUNIT INSIGHT CXR thresholds of 15,30, and 45, with 30 selected for analysis(19). The final study did not specify a DecXpert threshold(17).

The reference standards for TB diagnosis differed: one study used both culture-confirmed diagnosis and Xpert MTB/RIF(23). Two studies used only Xpert MTB/RIF(16,17) while the other studies relied solely on culture-confirmed diagnosis(17,18).

Study sensitivities at author preferred thresholds ranged from 0.73(95%CI: 0.61, 0.83)(16) to 1.00 (95%CI: 0.59, 1.00)(23). Specificities ranged from 0.60 (95%CI:0.53, 0.67)(19) to 0.88(0.84,0.91)(23). One study used two CAD software tools; CAD4TB and qXR, and showed identical diagnostic accuracy(18). The thresholds for CAD varied across studies; Table 2, S1 Appendix and S2 Appendix show details of each cut-point.

**Table 2:** Outcome Measures of the Included Studies.

| STUDY ID | Number of CXRs used for testing | Prevalence of Active TB % (95% CI) | Sensitivity % (95% CI) | Specificity % (95% CI) | Positive Predictive Value % (95% CI) | Negative Predictive Value % (95% CI) | Area Under the Curve (95% CI) |
| --- | --- | --- | --- | --- | --- | --- | --- |
| Habib 2020 | 694 | 10.7% (0.09, 0.1) | 73.0(61.4, 82.7) | 69.5(65.7, 73.1) | 22.2(19.2, 25.5) | 95.6 (93.7, 96.9) | 0.7(0.70, 0.75) |
| Khan 2020 | 270 | 23.3% (0.2, 0.3) | 98.4(91.5,100.0) | 73.4 (66.9,79.3) | 53.0 (47.3,58.6) | 99.4 (95.6, 99.9) | 0.9(0.8, 0.9) |
| Koesoemadinata 2018 | 346 | 2.0% (0.008, 0.04) | 100.0(59.0,100.0) | 87.9 (84.0,91.2) | 14.6 (11.4,18.5) | 100.0(98.8,100.0) | 0.9 (0.9, 1.0) |
| Nath 2024 | 299 | 34.8% (0.69, 0.82) | 88.5(80.7, 93.9) | 85.1(79.3, 89.8) | 76.0(69.2,81.7) | 93.3(89.0, 95.9) | 0.86(0.8, 0.9) |
| Tavaziva 2022 | 270 | 23.3% (0.2, 0.3) | 87.3 (76.5, 94.4) | 60.4 (53.4,67.1) | 40.2 (35.6,44.9) | 94.0 (89.0, 96.8) | 0.7 (0.7, 0.8) |

Figure 2 provided a forest plot stratified by the software used (CAD4TB, LUNIT Insight or DecXpert). For the 3 studies using the same software (CAD4TB) we reported results standardised to an approximate threshold of 60 (the mostly commonly used threshold for CAD) as shown by S1 Figure. Heterogeneity in results is evident, revealing variability in performance even after standardisation among the 3 CAD4TB studies, suggesting that factors beyond threshold setting affect accuracy.

**Figure 2.**
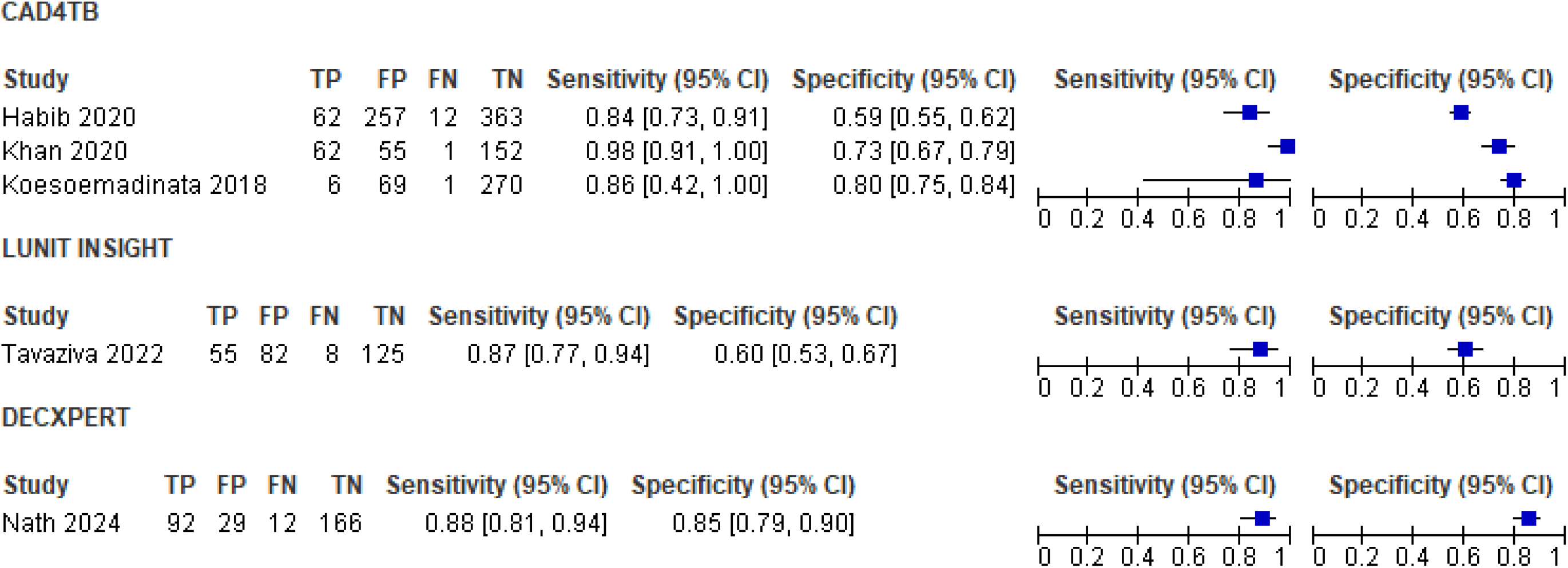
Forest plot of the 5 studies with their respective stratified CAD software. The Upper CAD4TB studies were standardised to the same approximate CAD threshold of 60 while the lower Tavaziva Study employed LUNIT INSIGHT at the given threshold of 30 whereas the DecXpert threshold employed by the Nath 2024 study did not explicitly specify the threshold.

The Area Under the Receiver Operating Characteristic Curve (AUC) varied from 0.7(95% CI:0.70, 0.75), to 0.9(95%CI:0.8,0.9) as shown by S8 Figure; indicating generally good to excellent diagnostic accuracy. One study that was conducted in a community setting, showed somewhat lower AUC compared to other studies taking place in diabetes treatment centres(16). False positive rates ranged from 0.24% to 30.5% at the author preferred thresholds whereas the false negative rates ranged from 0% to 3.2%.

No studies compared CAD software with other potential screening tests for TB disease (such as symptom screening, human reading of CXR) and likewise no studies compared the diagnostic accuracy of CAD among patients with diabetes compared to those without diabetes.

The quality appraisal of the included studies revealed overall good to excellent methodological quality, with 90% of the assessments indicating a low risk of bias across the four key domains as shown by figure 3 and S3 Figure. The flow and timing domain showed some variation, whereby 60% of studies showed low risk of bias across all domains, demonstrating excellent methodological quality (17–19) and 40% raised some concerns(18,19) in the flow and timing domain i.e. one study didn’t have a clear mention of patient flow details including the number of patients who were lost to follow-up or missed the CAD4TB index (16). In the Indonesian study most of the potential patients for inclusion had their DICOM (Digital Imaging and Communications Medicine) files accidentally deleted by the time of the study. Although many files were missing, they did not appear to be systematically missing since the deletion had occurred by accident(23).

**Figure 3.**
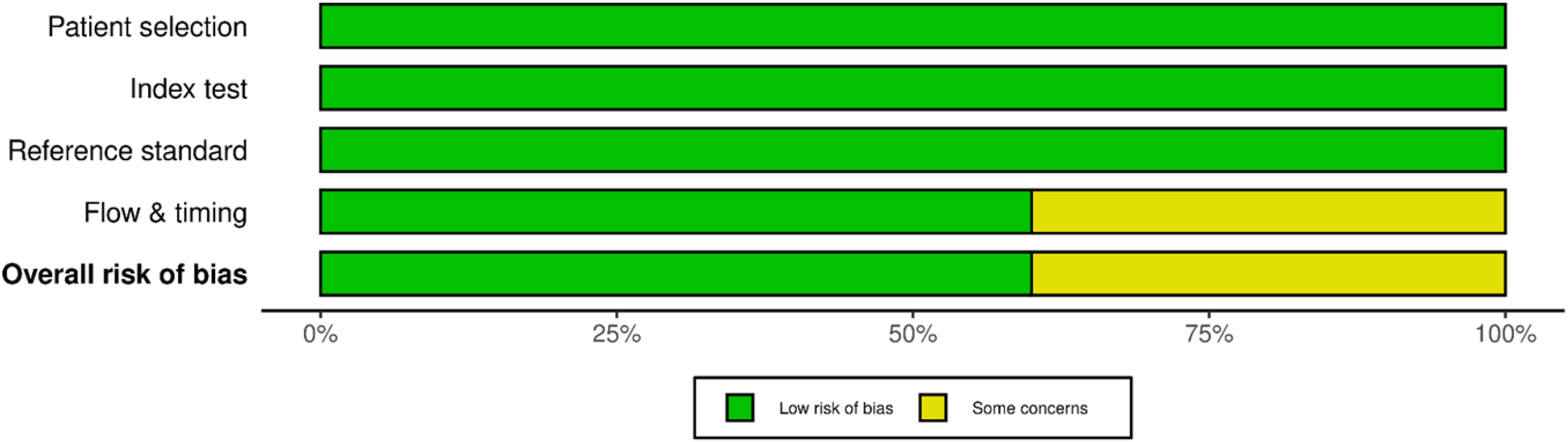
Traffic light methodological quality graph.

## DISCUSSION

This systematic review examined the accuracy of CAD in screening people with DM for active PTB. Our findings reveal a significant scarcity of data, with only five studies totalling 1,879 participants included. Notably, there was limited information on diabetes characteristics that could affect TB risk or diagnostic performance. This underscores a significant gap in existing literature regarding CAD performance in this high-risk group. The review highlighted the variability in CAD performance across the included studies, which utilised different software versions (CAD4TB, qXR, LUNIT INSIGHT and DecXpert). One small study used two different CAD software tools (CAD4TBv6.0 and qXR v2.0) on the same population, yielding identical results. Despite the limitations, the results suggest promising performance of CAD systems in this high-risk population with sensitivities ranging from 0.73 to 1.00, and specificities from 0.60 to 0.88 across different CAD software versions and settings.

The variability in CAD performance across studies highlights important considerations for its application in practice. Factors such as study settings, population demographics, TB prevalence, diabetes “severity” or comorbidities, and CAD software versions as well as threshold effects all potentially influenced the results. The apparent improved diagnostic accuracy in diabetes clinic studies compared to community settings suggests that disease severity and screening context may play crucial roles in CAD performance. However, the limited number of studies and their geographical concentration in Asian contexts (only three countries represented) underscore the need for caution in generalizing these findings to other populations or regions with different TB epidemiology.

The generally low false-negative rates at recommended thresholds are encouraging, suggesting that CAD systems may effectively minimize missed TB diagnoses in people with diabetes. However, the higher and more variable false positive rates observed across studies raise concerns about potential patient anxiety and resource allocation for additional TB testing. This variability emphasizes the importance of careful threshold selection in clinical practice, balancing sensitivity and specificity based on local resources and TB prevalence.

Our findings both align with and expand upon previous reviews of CAD in other population results. Our findings are consistent with a study conducted in 2019 which focused on the general population with sensitivity and specificity findings of (0.85 to 1.0) and (0.23 to 0.69) respectively (28) and the recent systematic review published in January 2024 which examined African Studies with mostly HIV patients revealing a sensitivity of (0.53 to 0.89) and a specificity of (0.56 to 0.98)(12). They all found generally high sensitivities with more variable specificities for different versions of CAD4TB and both did not perform meta-analyses due to methodological heterogeneity in the included studies.

However, caution should be exercised when interpreting these findings due to the relatively small number of studies among people with DM and lack of direct comparisons of different population groups in any one study.

This review has several strengths, including its focus on CAD for TB screening specifically in people living with diabetes, addressing a critical gap in research. It employed a very comprehensive literature search, ensuring inclusivity of studies and minimising publication bias. The methodological quality of included studies was rated good to excellent, bolstering the reliability of findings However, limitations included the limited number of studies (only five) thus restricting generalizability. Variability in reference standards and thresholds among studies complicates comparisons, and the inability to compare CAD performance between people with diabetes from the general population highlighting a significant research gap. Furthermore, all included studies were conducted in Asian contexts (only 3 different countries), potentially limiting the applicability of findings to regions with different TB epidemiology.

In conclusion, while CAD shows promise for improving TB screening efficiency among people with diabetes, particularly in resource limited settings, further research is imperative. Future studies should directly compare CAD performance across different risk groups, explore the impact of diabetes-related factors on CAD accuracy, and evaluate cost-effectiveness in various clinical contexts. Standardization of reference standards and CAD thresholds would enhance comparability across studies. Additionally, qualitative research into patient and provider perspectives on CAD implementation could inform strategies to overcome potential barriers to adoption. As CAD technology continues to evolve, its potential role in managing other pulmonary complications associated with diabetes should also be explored, potentially expanding its clinical utility in this high-risk population.

## Data Availability

All relevant data are within the manuscript and supporting information files.

## Acknowledgements

Ms Jane Falconer helped with refining the search strategy of this project.1

## Funding

1. Reagan (EDR) was supported primarily by a Chevening scholarship as part of his Master’s in Public Health in London.
2. JAC, RvC and LEM were supported by EDCTP funded PROTID study Preventive Treatment of Latent Tuberculosis Infection in People with Diabetes Mellitus (PROTID) study, which is part of the EDCTP2 program supported by the European Union [grant number RIA2018CO-2514-PROTID]

## Notes

### Competing Interest Statement

The authors have declared no competing interest.

### Funding Statement

Yes

### Author Declarations

It was a systematic review that really did not require any ethical approval (N/A)

